# Examining stability of dietary patterns across time using repeated measures of dietary intake: Results from Black Women’s Health Study

**DOI:** 10.64898/2026.08.08.26360017

**Authors:** Briana Joy K. Stephenson, Xuzhi Wang, Walter C. Willett, Jessica L. Petrick, Julie R. Palmer

## Abstract

**Background:** Many epidemiological studies rely on dietary exposures taken from baseline only. This limits our understanding of diet-disease associations because it requires assuming a level of temporal stability, either by individuals or dietary pattern composition.

**Objectives:** This study aimed to evaluate these analytic assumptions of pattern structure consistency and baseline adherence using a cohort of US Black women with repeated measures of food frequency questionnaires (FFQ).

**Methods:** Data from 6151 Black women aged 21-69 from the Black Women’s Health Study with complete FFQ data in 1995, 2001, 2013, and 2021 were evaluated for temporal stability. Baseline dietary patterns were derived using an overfitted latent class model. Parameter estimates from the baseline model were then applied to subsequent waves to track individual transitions between existing patterns. Dietary patterns were also derived at each time point using an overfitted latent class model and assessed for changes in pattern composition over time.

**Results:** Five baseline dietary patterns were identified in 1995. Only 18% of participants remained in the same baseline dietary pattern across all four time points, while all others transitioned to a different baseline-derived pattern. Dietary patterns derived independently at subsequent time points, yielded a different number of dietary patterns at each time point (2001: 6 patterns, 2013: 5 patterns, 2021: 4 patterns). Correlation strength of subsequent derived patterns and baseline patterns significantly weakened in strength after 2001 (40% pairings > 0.5), with no patterns correlated greater than 0.5 in 2021.

**Conclusion:** Prospective studies that rely on baseline dietary exposure data cannot assume stability of pattern composition or individual pattern adherence over time, as it ignores changes in dietary habits and may bias our understanding of the diet-disease pathway.

## INTRODUCTION

Diet assessments, such as the food frequency questionnaire (FFQ), have long been used to examine dietary consumption behaviors in target populations, and poor diet has long been identified as a major contributor to the onset and progression of cardiovascular disease (1,2). Many epidemiological studies have collected dietary exposure information at baseline, but few prospective studies continue to make subsequent collections of dietary information due to budgetary constraints, participant burden, and other logistical concerns. As a result, dietary intake is often considered a baseline exposure, while other clinical markers (e.g. BMI, blood pressure, glucose level, cholesterol) are often collected repeatedly for the duration of the longitudinal study. This approach relies on the assumption that ranking of dietary intake remains consistent/stable over time. While minimal changes have been observed in diet in the span of a few years, more substantial changes are possible over longer periods of time as individuals experience changes in their lifestyle, cultural and social environment, financial resources, or geography (3). Yet, few studies have the capability to examine how consistent these habits are maintained over time. Cohorts that do are primarily focused on predominantly White or pediatric populations. This raises a concern that true dietary intakes of populations at greater risk of suboptimal diets may be masked by the assumptions that temporal patterns of intake for a majority demographic group apply to important subgroups. In nationally representative studies, Black women have been frequently cited to be at greater risk of suboptimal diets compared to other subgroups(4–10).

The Black Women’s Health Study is a prospective cohort study that has been examining the causes, needs, and factors associated with health and well-being of Black women in the United States since 1995. This is one of the few population-based studies centered solely on Black women that includes repeated measures dietary data.

Empirically-based dietary patterns are considered a comprehensive exposure measurement to examine diet-disease relationships (11,12). These patterns can be empirically derived using a latent class framework. (13–16). However, applied in a longitudinal setting, methods that have been applied to dietary data, such as group-based trajectory model and growth mixture models, are constrained by the assumption that patterns are either a) defined at baseline and remain unchanged over time, or b) individuals within a pattern follow the same trajectory changes over time (17).

This study aims to challenge those two assumptions by investigating the temporal stability of dietary patterns over time by (1) identifying baseline dietary patterns and using their parameters to classify participants at later waves, assessing whether individuals remain in or transition between existing patterns, and (2) deriving dietary patterns independently at each time point to assess whether pattern composition itself changed over time. Using dietary intake data from the Black Women’s Health Study, we plan to empirically derive dietary patterns of participants and assess pattern structure consistency and baseline adherence from 1995 to 2021.

## METHODS

### Data Source

The Black Women’s Health Study (BWHS) is a prospective cohort study following 59,000 Black American women, aged 21-69 years old in 1995. These participants were enrolled in 1995 through a health questionnaire mailed to subscribers to Essence Magazine (publication targeted for Black American women). In 1995, 2001, 2013, and 2021, participants were asked to complete a semi-quantitative food frequency questionnaire (FFQ). A total of 68 food and beverage items were queried in 1995. This list was expanded to 85 items in subsequent collection years. The four FFQs were harmonized across the four cycles for this analysis. Six foods were deleted that did not appear in the 1995 and 2001 questionnaires (soy, soy burger, pizza, water, tofu, fat added on vegetables). Several foods were merged to have consistency across all four cycles, this includes: Oranges and grapefruit; bacon and sausage; regular and low-fat salad dressing or mayo; regular and low-fat ice cream; dark fish, other fish, and shellfish. Chicken or turkey and chicken pot pie were combined in 1995 and harmonized with future cycles where we combined dishes with poultry and other poultry. Similarly, spaghetti, lasagna, and other pasta from 1995 were harmonized with future cycles where we combined spaghetti and macaroni and cheese. After all foods were deleted or merged, 65 food items were included for analysis. Each food item was queried based on 10 frequency responses ranging from less than once a month to at least twice per day (food) or at least 6 times per day (beverage), as well as portion size defined for each level (e.g. medium serving = ½ cup of green beans). To capture both queries, frequency of consumption reported was multiplied by the reported USDA portion weight size of that item and multiplied by a factor based on the reported size of consumption.

Medium portion size used a factor of 1. Small portion size used a factor of 0.5. Large portion size used a factor of 1.5. Super portion size, which was included in 2001, used a factor of 2. After the three terms were calculated, the relative frequencies of consumption were recalculated based on the amount of consumption. For example, if consumed once a week the value was multiplied by a factor of 1/7. Levels of frequency were collapsed to reduce sparse levels of consumption. Number of levels for each food item ranged from 2 (less than once a month vs more than once a month) to 7 levels (less than once a month to at least 5 times per week). **Supplementary Table 1** provides a list of the 65 food items analyzed, with the number of consumption levels, and observed frequencies at each analyzed consumption level. Baseline demographic information of participants was included for descriptive purposes including age, education, occupation, marital status, parent care, childcare responsibilities, household size, physical activity, and smoking status. Participants who had complete dietary data at all four time points were included for analysis (n=7281). This number reflects the fact that the 2013 FFQ was appended to the end of the online version of the 2013 BWHS follow-up questionnaire, but was not included in the paper version, which, at that time, was completed by a majority of respondents. Participants who were pregnant at the time of observation (e.g. 1995, 2001) were excluded from analysis. After inclusion/exclusion criteria were performed, a total of 6151 participants were included for analysis. Alternative Healthy Eating Index 2010 (AHEI-2010) is a diet quality score created to measure adherence to a set of foods and nutrients that are associated with a decreased risk of chronic disease(18). This score was calculated for each participant at each time point to best contextualize the dietary patterns derived and provide a standardized measure of comparison.

### Overfitted finite Mixture Model

Dietary patterns were derived using an overfitted latent class model at baseline (1995) (19). Structured like the standard latent class model first introduced by Lazarsfeld (20), the overfitted latent class model does not assume to know the number of dietary patterns *a priori*. Instead, the latent class model is fit with an oversized number of dietary patterns and estimated using an MCMC Gibbs Sampling algorithm. At the end of the sampling algorithm, the empty clusters will drop out and the filled clusters will remain. A filled cluster is defined as a cluster containing at least 5% of the sampled population. Participants that have not yet been assigned to one of the nonempty filled clusters, are assigned using the posterior estimate summaries (e.g. median) of each of the parameters and assigning that participant to the cluster with maximum probability of assignment.

To identify dietary patterns at each time point, separate overfitted mixture models were applied to consumption data collected in 1995, 2001, 2013, and 2021. Pattern differences were assessed at separate time points by comparing the level of consumption for each food item that had the highest posterior probability (i.e. mode) within each derived pattern. We refer to this as the modal consumption pattern. Two approaches were implemented to analyze transition of diet over time. Approach 1 applied the parameter estimates from the overfitted mixture model derived in 1995 to the observed dietary data collected in 2001, 2013, and 2021. Participants were assigned to the dietary pattern with the highest probability of assignment. Pattern stability was evaluated against the baseline dietary pattern using Pearson’s correlation coefficients for the modal consumption pattern of each model. Transition plots were generated to illustrate if or where participants assigned to one pattern moved to a new pattern in the subsequent time point. Approach 2 utilized the separate overfitted mixture model results from the four time points. Here, dietary patterns at the different time points were aligned as close as possible based off shared characteristics in their consumption distributions. In this approach, the number of derived patterns may differ for different time points. Transition plots were then generated to illustrate where participants were re-assigned for subsequent time points.

## RESULTS

Appendix A (**Supplementary Table 1)** provides a descriptive table of the consumption distribution of the 65 food items for the BWHS sample included for analysis. Consumption frequencies showed similarities in 1995 and 2001. Other similar frequencies were seen from 2013 and 2021. Greater shifts were seen from 2001 to 2013. Overall, foods such as fruit drinks, soda, fries/potatoes, other cereal, pasta, bread, and hamburgers saw a general decrease in consumption frequency over time. Foods such as spinach, other vegetables, and tomatoes saw a general increase in consumption frequency over time. **Supplementary Table 2** provides a summary of AHEI2010 scores and subcomponents across each of the four time cycles. Overall, diet quality of participants with complete diet data have improved steadily since baseline. Main contributors to this improvement, were decreasing consumption of sugary sweetened beverages and sodium, and increasing consumption of omega-3 fatty acids.

### Baseline Dietary patterns

At baseline (1995), five dietary patterns were identified. Figure 1 provides an illustration of the derived pattern by mode of consumption (Figure 1A) and a complete distribution of derived 1995 patterns (Figure 1B). Participants assigned to pattern 1, with a mean AHEI2010 score of 39.3 (SD: 7.9), had the lowest diversity of foods, with only ten foods favoring a positive consumption: spaghetti, white and dark bread (at least 1x/week); salad dressing, poultry, rice, salad, and broccoli (at least 2-3x/month).

**Figure 1:**
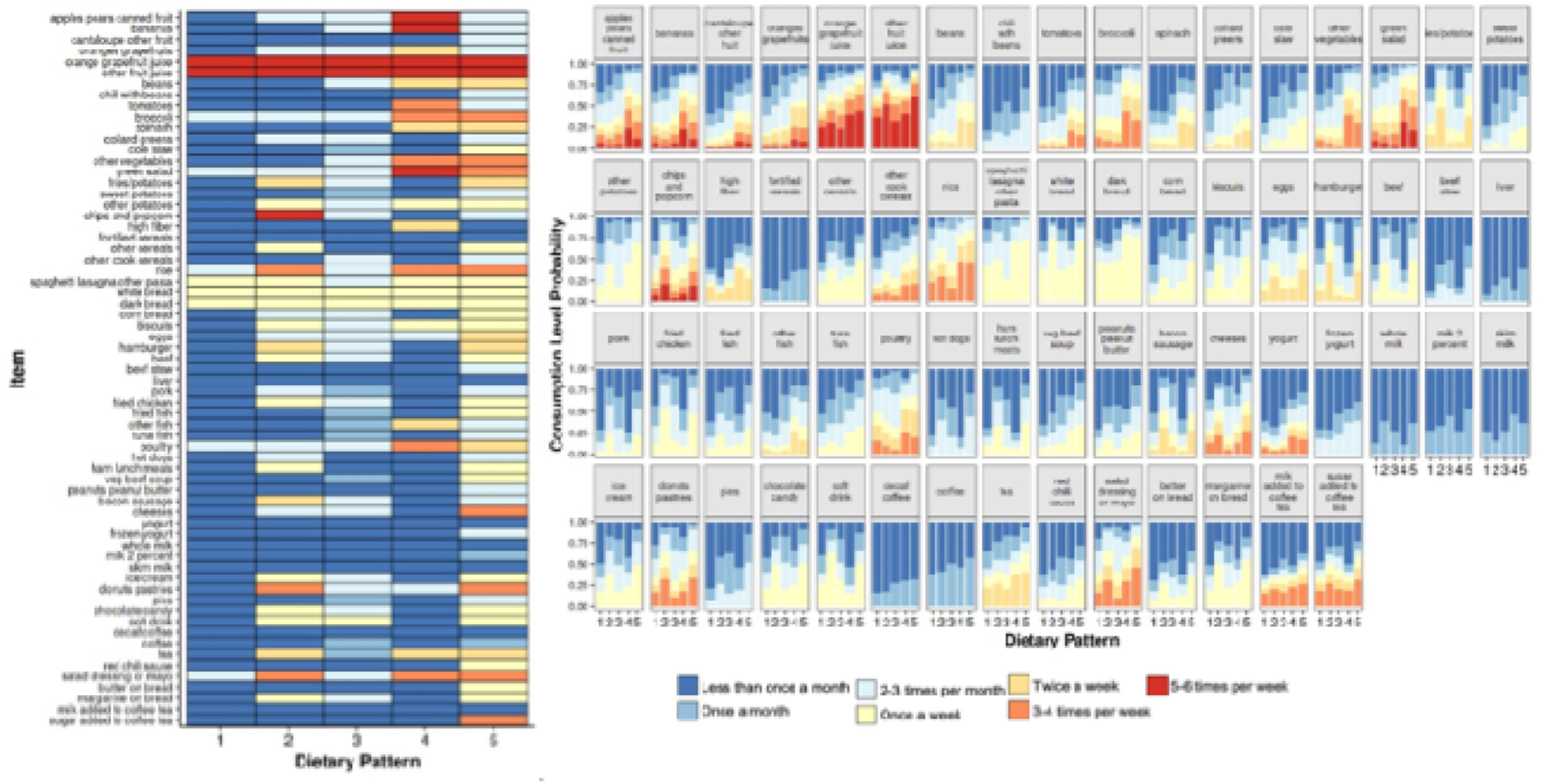
Modal consumption patterns derived from 1995 (left). Full dietary pattern distribution, where length of bar denotes the magnitude of the consumption level distinguished by colors (right).

Participants assigned to pattern 2, with the lowest mean AHEI2010 score of 30.4 (SD: 6.8), favored higher consumption frequencies of sweets and snacks such as chips/popcorn (5-6x/week); donuts/pastries (3-4x/week); and hamburger, fries/potatoes, and bacon/sausage favored consumption (2x/week). Participants assigned to pattern 3, with a mean AHEI2010 score of 39.6 (SD:6.5), had a moderate level of consumption (at least 2-3 times per month) over many of the foods queried (43 foods). For example, they favored consumption of leafy greens (spinach, collard greens, salad), potatoes, rice, pasta, breads, eggs, burger, fried meats, fish, and dessert foods (ice cream, donuts, chocolate) 2-3 times a month. Participants assigned to pattern 4, with the highest mean AHEI2010 score of 47.3 (SD: 8.3), favored more frequent consumption of prudent items such as fruit, juice, and spinach (5-6x/week); green salad, tomatoes, broccoli, other vegetables, rice, poultry (3-4x/week); high fiber and other fish (2x/week). Participants assigned to pattern 5, with a mean AHEI2010 score of 37.4 (SD: 8.1), had the largest variety of foods favored to be consumed of all the groups, with higher consumption frequencies for juice (5-6x/week); broccoli, other vegetables, green salad, rice (3-4x/week); beans, fries/potatoes, eggs, poultry (2x/week), and sweets like pies, donut, ice cream and chocolate (at least 1x/week). These patterns were roughly similar in size. Pattern 4 had the largest proportion of participants assigned (n_4_=1626, 26%). Pattern 1 had the smallest proportion of participants assigned (n_1_=1020, 17%).

Fitting the 2001, 2013, 2021 data to the model parameters of the baseline model shifted participants to different patterns in subsequent years. **Figure 2** shows the transition of participants being re-assigned to different patterns in subsequent waves. After 1995, about half of the participants were assigned to the same baseline pattern (Pattern 1=49%, Pattern 2=40%, Pattern 3=48%, Pattern 4=59%, Pattern 5=58%). However, after 2001, most participants transitioned to a new pattern in 2013, and again in 2021. Participants assigned to pattern 4 (highest mean AHEI score) at each time point were the least likely to shift to a different pattern at a subsequent time point. Across all four time points only 18% of participants remained in the same baseline diet pattern across all time waves (Pattern 1=20%, Pattern 2=6%, Pattern 3=13%, Pattern 4=32%, Pattern 5=16%).

**Figure 2:**
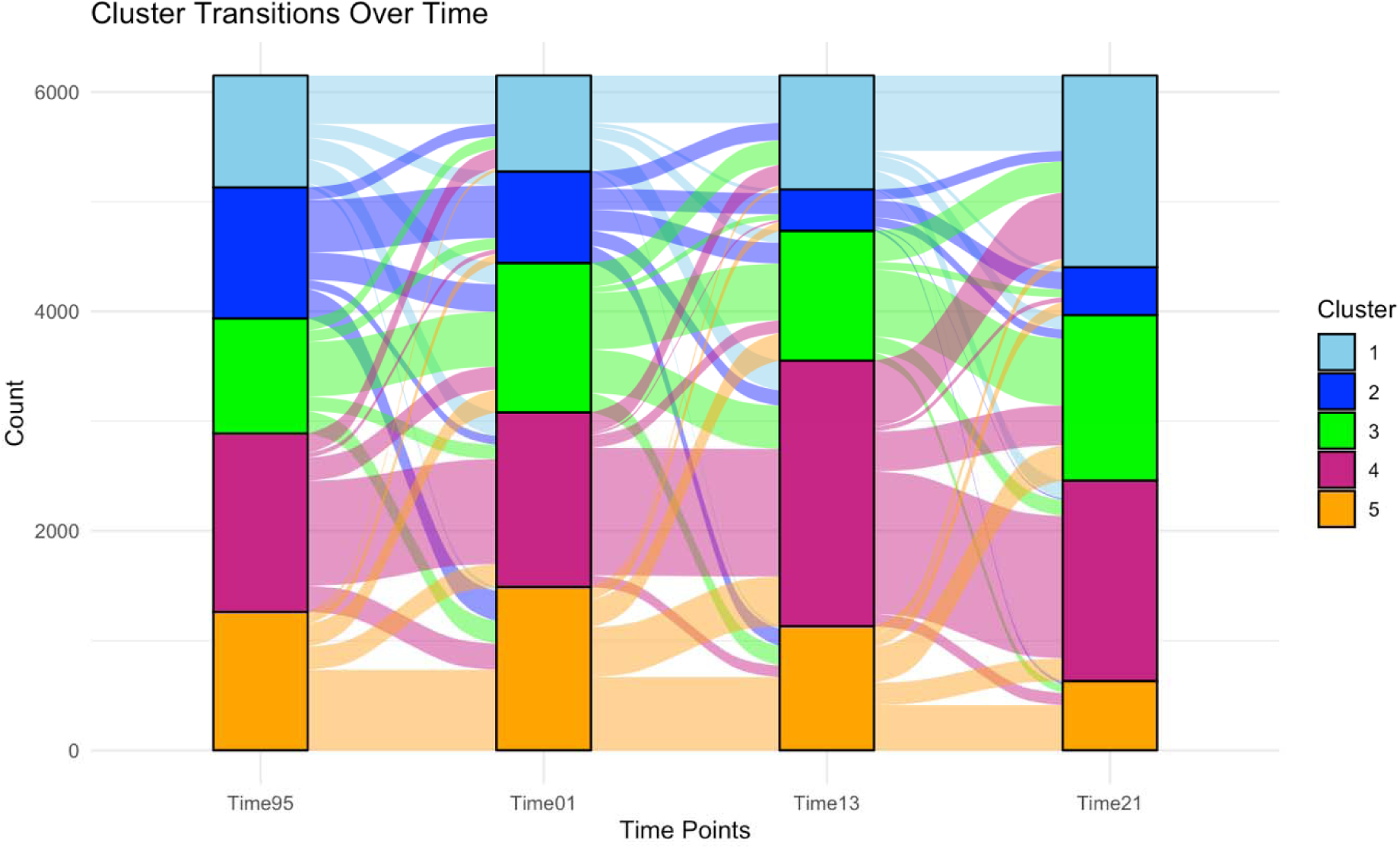
Dietary pattern assignment transitions over time. Thicker bands denote larger proportion of participants who transitioned from one dietary pattern to the same new dietary pattern at the subsequent time point

### Time-Independent Dietary Patterns

When models were run separately for each time point, a different number of dietary patterns were identified with varying characteristics. Six dietary patterns were identified in 2001. Five dietary patterns were identified in 2013. Four dietary patterns were identified in 2021. Four foods (fortified cereal, liver, whole milk, decaffeinated coffee) favored the same levels of consumption (less than once a month) across all dietary patterns and four time points.

**Figure 3** provides modal consumption patterns of the 65 foods queried analyzed at each time separately. **Supplementary Figures 1-4** provide a more comprehensive examination of each pattern illustrating the probability of consumption at each potential level of each food. Details about each pattern for each time point are also detailed in **Appendix B**. None of the baseline dietary patterns were reproduced at later time waves. However, several patterns shared similar modes of consumption, which may indicate the model’s tendency to cluster participants together based on the shared consumption of a few similar foods.

**Figure 3:**
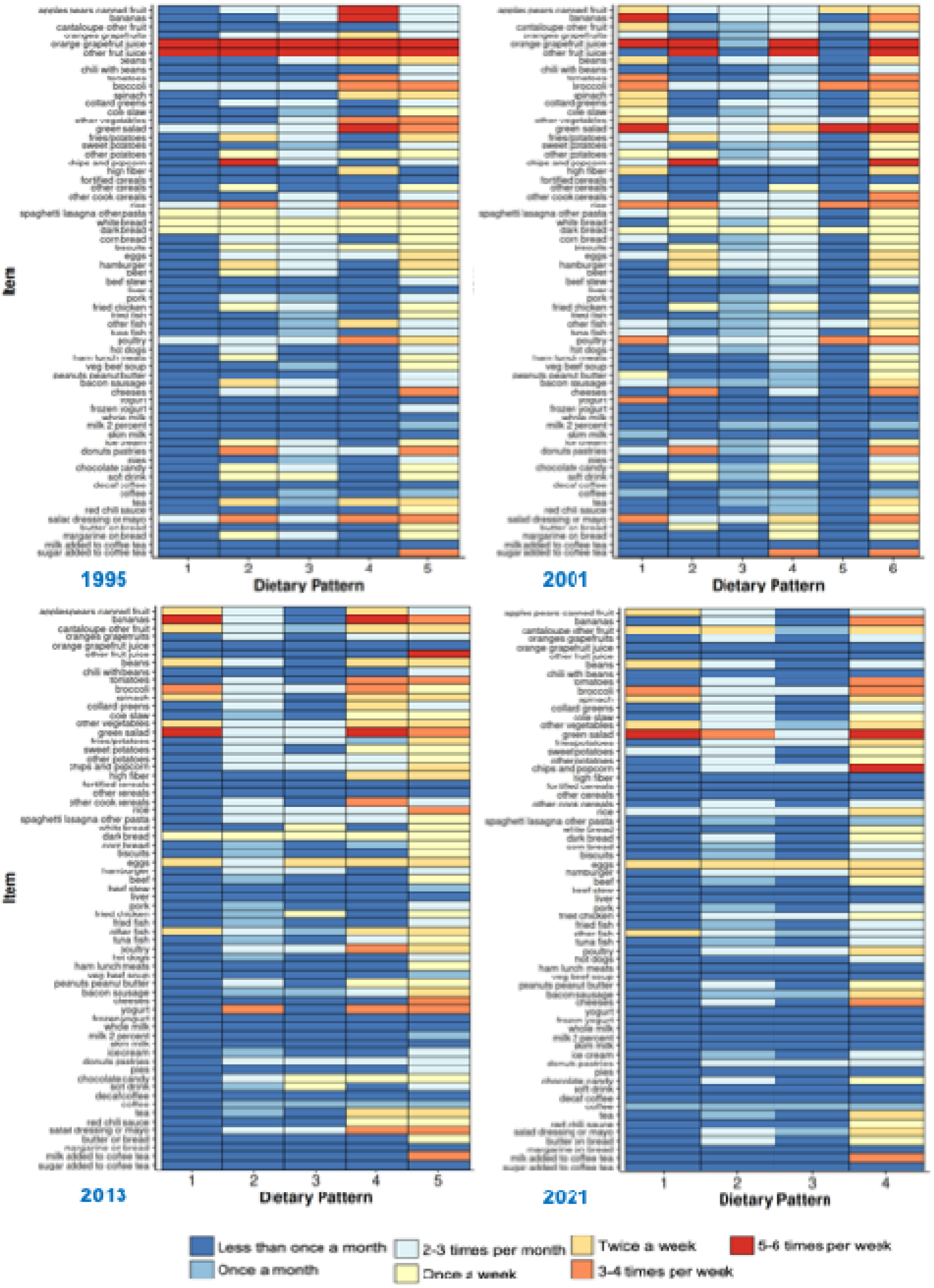
Heatmap illustrating the dietary patterns derived at each time point (1995, 2001, 2013, 2021). Pattern is determined based on the consumption level with the highest probability for participants assigned to that respective cluster.

**Supplementary Figure 5** provides correlation plots to understand how patterns described by the mode of consumption varied from the baseline consumption patterns. The patterns derived in 2001 had the highly correlated patterns (Supplementary Figure 5A). Twelve parings had correlations greater than or equal to 0.5. The strongest correlation in modal consumption patterns was seen with P2^1995^ and P2^2001^ (0.72) and P3^1995^ and P3^2001^(0.69). In 2013, only three pairings had correlations greater than 0.5 (P3^1995^-P3^2001^= P4^1995^-P4^2001^=0.54; P4^1995^-P1^2001^= =0.53). In 2021, no patterns had a correlation stronger than 0.5 with the baseline pattern, and many pairings were not statistically significant.

**Figure 4** provides an illustration of how participants assigned to one pattern at one time point transitioned to a different pattern at another time point. While no pattern was identical, dietary patterns that shared broadly similar characteristics were assigned the same color across the four time points. Light blue pattern shared similar consumptions of fruit, leafy greens, and eggs. Royal blue shared an overall low variety of foods consumed. Green pattern had a medium to high variety and shared similar consumption habits of meats, potatoes, and dairy. Purple pattern had a high variety of foods consumed but at an overall low frequency consumption. Orange pattern had a moderate variety of foods consumed but lower frequencies of consumption for fruit, eggs, and vegetables. The thicker the band the larger the proportion of participants who were assigned to the same sequence of patterns at adjacent time points. There were 545 unique transitions between the four time points. As illustrated previously, P4^1995^,P1^2001^,P4^2013^,P1^2021^ shared some pattern similarities. About 4 percent of the cohort shared this exact transition sequence (n=234, 3.8%). These patterns had the highest mean AHEI2010 score in their respective years, reflecting the same level of positive consumption of beans (at least twice a week), broccoli (twice a week), green salad (at least 5-6 times a week) across all four time points. Meats and poultry had higher consumption levels in 1995 and 2001, whereas eggs and other fish favored higher consumption in 2013 and 2021. Foods such as rice and tomatoes favored consumptions at 3-4 times per week until 2021 where the P1^2021^ pattern favored 2-3 times per month and none/less than once month, respectively.

**Figure 4:**
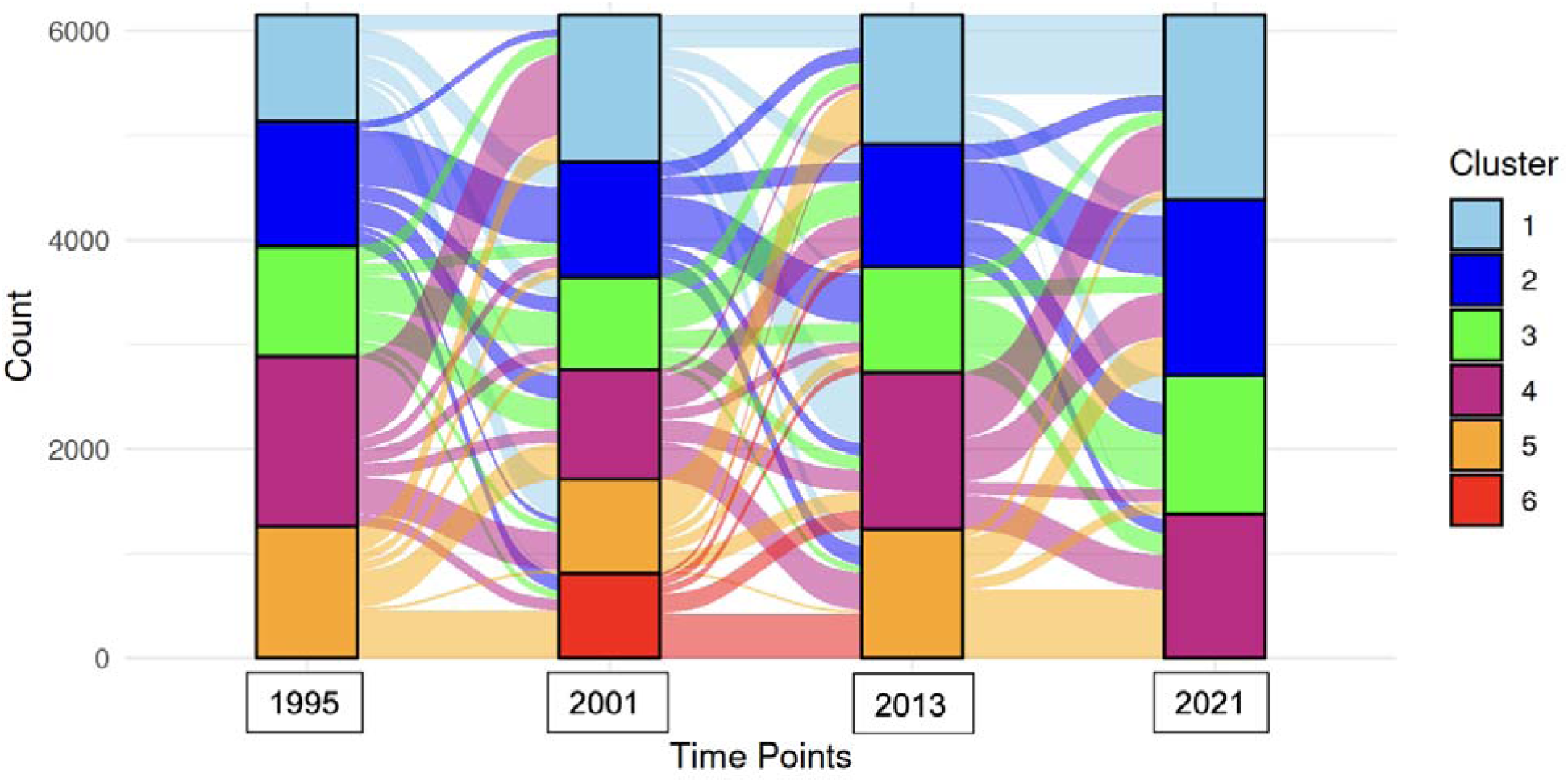
Transition plot illustrating the relative proportion of participants who transitioned from one pattern (denoted by color) to a new pattern in the subsequent time point. Wider bands denote larger proportion of participants transitioned together.

Comparing across all four years, P5^1995^, P6^2001^, P5^2013^, P4^2021^ had the highest level of diversity of foods with a favored positive consumption. These patterns had the lowest AHEI2010 mean scores in years 1995, 2013, and 2021. At least a monthly level of consumption was found in almost all food groups except for dairy products (milk, yogurt). Higher levels of consumption were favored in foods such as juice (before 2021), cheese, rice, salad dressing, tomatoes, broccoli, bacon/sausage, tea, and chocolate. However, only 3% (n=194) of participants shared this exact transition sequence across the four time waves.

## DISCUSSION

Our analysis evaluated the stability of dietary patterns over time since baseline for participants in the Black Women’s Health Study. At baseline, five dietary patterns were derived. Under the assumption that these patterns remained stable over time, we showed that most participants transitioned to different dietary patterns at later time points. These transitions varied from pattern to pattern and from time point to time point. When patterns were derived independently at each time point, we identified a different number of patterns at each time point with different food consumption patterns composing of these patterns. These results suggest that when following participants across time, analysis should not assume “pattern-level” stability, where behavior patterns identified at baseline remain consistent through the study period, nor should they assume “individual level stability”, where participants who share consumption behaviors at baseline also share the same changes in consumption behaviors with those same clustered participants over time.

This is not the first study to examine the stability of dietary patterns over time, but it is the first to explicitly focus on the dietary patterns of Black women over time. Prior studies, which focused on pediatric, European, Asian, Canadian, or pregnant populations, have evaluated diet stability but differed in approach. We provide a brief overview of prior works detailing their strengths and limitations in **Supplementary Table 2**. These approaches often examined variation in a univariate measure (e.g. adherence score, factor loading) that summarized trends in stability of the overall population, but failed to capture individual variation of foods and participants over time (10,21). Our analysis is one of the first to examine this stability directly providing a more comprehensive and nuanced view of when and how these patterns shift in composition and assignment over time. Further, our examination of diet pattern stability in Black women participants of the Black Women’s Health Study provided an additional layer of novelty to the literature as this population is severely understudied in the nutrition literature. Within the BWHS literature, diet intake was assessed in 1995, 2001, 2013, and 2021, but analyses have published based on1995 and 2001 data only. Two BWHS studies used a factor analysis to derive dietary patterns, grouping foods together that share similar variation (22,23). However, as previously noted, this dimension reduction technique limits the characterization of these patterns with a loss of individual food level information.

Despite its strengths, this analysis does still have limitations. Firstly, each of the approaches discussed and implemented are population specific. Different populations yield different model estimates, and consequently different dietary patterns. Given that this is the first study to highlight specifically the dietary consumption behaviors of US Black women, we are unable to confirm if the lack of pattern stability is specific to this study population, which is self-selected and demographically more educated with relatively high study response rates, or generalizable to the overall US Black women population, as well as other populations that have previously been analyzed.

Researchers should evaluate their study populations for individual and pattern level stability before considering which assumptions are appropriate when selecting their analytic method. Secondly, our results rely on repeated measures of self-reported food frequency questionnaires. These types of assessment tools leave room for measurement error and recall bias. While no method is perfect, the BWHS FFQ has previously shown to have moderate-to-strong validity, which makes this still a reliable instrument to understand dietary consumption behaviors in this population(24,25). Third, our analysis relied on complete dietary intake data. Consequently, our results only reflect a fraction of the total study population. Participants not included in this analysis might differ in their dietary stability over time. Therefore, our results may be over or underestimating the true stability of the population’s patterns over time. Imputation methods could address some of the missingness but was not used in this study to avoid additional bias and error in our population-based results. Further research is needed to allow these clustering models to handle missingness across time points so that results can be generalizable to the full study population.

The role of diet and health is a dynamic and complex process. Epidemiological studies that limit dietary exposure observed at baseline measure associations with health outcomes at later time points may not be telling the full story. Misclassifying dietary patterns to individuals in prospective epidemiological studies risk will generally bias the association of diet with a given health outcome towards the null. Our analysis confirms that baseline diet may not be a suitable proxy for long-term dietary exposure. Methods are needed to properly account for time-varying pattern membership and individual variability of diet. Recent methods have been developed in genomic literature that are able to analyze repeated measures of multivariate, high-dimensional exposures(26–30). However, these methods are still limited in scalability (handle the large number of foods in a dataset), dependencies (handle correlated foods that are commonly consumed together), and data structure (handle flexibility of different data types as well as sparsity and skewness typically seen in foods that are rarely or commonly consumed). Further research is needed to overcome these data challenges and provide a more accurate examination of the impact of dietary exposure pattern changes over time.

Until more improved methods are developed and applied, interventions and policies that are based on dietary patterns should remain as current and up to date as feasible so that guidelines can be adapted to sustain the full life course.

## Disclaimers

N/A.

## Abbreviations

BWHS: Black Women’s Health Study
FFQ: Food Frequency Questionnaire

## Data Availability Statement

Data used in the analysis and reporting of this study are not publicly available. Data can be made available upon request and approval of a concept proposal on the study website: https://www.bu.edu/bwhs/for-researchers/data-requests/

## REFERENCES

1. The Burden of Cardiovascular Diseases Among US States, 1990-2016 | Cardiology | JAMA Cardiology | JAMA Network [Internet]. [cited 2026 Mar 24]. Available from: https://jamanetwork.com/journals/jamacardiology/fullarticle/2678113

2. US Burden of Disease Collaborators, Mokdad AH, Ballestros K, Echko M, Glenn S, Olsen HE, Mullany E, Lee A, Khan AR, Ahmadi A, et al. The State of US Health, 1990-2016: Burden of Diseases, Injuries, and Risk Factors Among US States. JAMA 2018;319:1444–72.

3. Weismayer C, Anderson JG, Wolk A. Changes in the stability of dietary patterns in a study of middle-aged Swedish women. J Nutr 2006;136:1582–7.

4. Richards Adams IK, Figueroa W, Hatsu I, Odei JB, Sotos-Prieto M, Leson S, Huling J, Joseph JJ. An Examination of Demographic and Psychosocial Factors, Barriers to Healthy Eating, and Diet Quality Among African American Adults. Nutrients [Internet] 2019 [cited 2026 Apr 28];11:519. Available from: https://pmc.ncbi.nlm.nih.gov/articles/PMC6470798/

5. Molitor F, Doerr C. Diet Quality Differs by Race/Ethnicity Among Mothers and Their Children from Supplemental Nutrition Assistance Program–Education Households. Health Equity [Internet] 2021 [cited 2026 Apr 28];5:633–6. Available from: https://pmc.ncbi.nlm.nih.gov/articles/PMC8665816/

6. Kirkpatrick SI, Dodd KW, Reedy J, Krebs-Smith SM. Income and race/ethnicity are associated with adherence to food-based dietary guidance among U.S. adults and children. J Acad Nutr Diet [Internet] 2012 [cited 2026 Apr 28];112:624–635.e6. Available from: https://pmc.ncbi.nlm.nih.gov/articles/PMC3775640/

7. Nguyen X-MT, Li Y, Whitbourne SB, Djousse L, Wang DD, Ivey K, Willett WC, Gaziano JM, Cho K, Hu FB. Racial and Ethnic Disparities in Dietary Intake and Quality Among United States Veterans. Curr Dev Nutr [Internet] 2024 [cited 2026 Apr 28];8:104461. Available from: https://pmc.ncbi.nlm.nih.gov/articles/PMC11530779/

8. Parker HW, Tovar A, McCurdy K, Vadiveloo M. Socio-economic and racial prenatal diet quality disparities in a national US sample. Public Health Nutr [Internet] [cited 2026 Apr 28];23:894–903. Available from: https://pmc.ncbi.nlm.nih.gov/articles/PMC10200547/

9. Stephenson BJK, Willett WC. Racial and ethnic heterogeneity in diets of low-income adult females in the United States: results from National Health and Nutrition Examination Surveys from 2011 to 2018. Am J Clin Nutr [Internet] 2023 [cited 2026 Apr 28];117:625–34. Available from: https://pmc.ncbi.nlm.nih.gov/articles/PMC10315405/

10. Tao M-H, Liu J-L, Nguyen U-SDT. Trends in Diet Quality by Race/Ethnicity among Adults in the United States for 2011-2018. Nutrients 2022;14:4178.

11. Hu FB, Rimm EB, Stampfer MJ, Ascherio A, Spiegelman D, Willett WC. Prospective study of major dietary patterns and risk of coronary heart disease in men. The American Journal of Clinical Nutrition [Internet] 2000 [cited 2026 Apr 9];72:912–21. Available from: https://linkinghub.elsevier.com/retrieve/pii/S0002916523068004

12. Schulze MB, Martínez-González MA, Fung TT, Lichtenstein AH, Forouhi NG. Food based dietary patterns and chronic disease prevention. BMJ [Internet] 2018 [cited 2026 Apr 9];k2396. Available from: https://www.bmj.com/lookup/doi/10.1136/bmj.k2396

13. Sotres-Alvarez D, Herring AH, Siega-Riz AM. Latent Class Analysis Is Useful to Classify Pregnant Women into Dietary Patterns123. J Nutr [Internet] 2010 [cited 2026 Apr 28];140:2253–9. Available from: https://pmc.ncbi.nlm.nih.gov/articles/PMC2981007/

14. Graf S, Cecchini M. Identifying patterns of unhealthy diet and physical activity in four countries of the Americas: a latent class analysis. Rev Panam Salud Publica [Internet] 2018 [cited 2026 Apr 28];42:e56. Available from: https://pmc.ncbi.nlm.nih.gov/articles/PMC6385803/

15. Stephenson BJK, Willett WC. Racial and ethnic heterogeneity in diets of low-income adult females in the United States: results from National Health and Nutrition Examination Surveys from 2011 to 2018. Am J Clin Nutr 2023;117:625–34.

16. Park JH, Kim JY, Kim SH, Kim JH, Park YM, Yeom HS. A latent class analysis of dietary behaviours associated with metabolic syndrome: a retrospective observational cross-sectional study. Nutr J [Internet] 2020 [cited 2026 Apr 28];19:116. Available from: https://pmc.ncbi.nlm.nih.gov/articles/PMC7568389/

17. Dalrymple KV, Vogel C, Godfrey KM, Baird J, Hanson MA, Cooper C, Inskip HM, Crozier SR. Evaluation and interpretation of latent class modelling strategies to characterise dietary trajectories across early life: a longitudinal study from the Southampton Women’s Survey. Br J Nutr [Internet] [cited 2026 Apr 28];129:1945–54. Available from: https://pmc.ncbi.nlm.nih.gov/articles/PMC10167664/

18. McCullough ML, Feskanich D, Stampfer MJ, Giovannucci EL, Rimm EB, Hu FB, Spiegelman D, Hunter DJ, Colditz GA, Willett WC. Diet quality and major chronic disease risk in men and women: moving toward improved dietary guidance. The American Journal of Clinical Nutrition [Internet] 2002 [cited 2026 Aug 3];76:1261–71. Available from: https://linkinghub.elsevier.com/retrieve/pii/S0002916523060574

19. Van Havre Z, White N, Rousseau J, Mengersen K. Overfitting Bayesian Mixture Models with an Unknown Number of Components. Chen CWS, editor. PLoS ONE [Internet] 2015 [cited 2026 Jan 5];10:e0131739. Available from: https://dx.plos.org/10.1371/journal.pone.0131739

20. Lazarsfeld PF, Henry NW. Latent Structure Analysis. Houghton, Mifflin; 1968.

21. Gasser CE, Kerr JA, Mensah FK, Wake M. Stability and change in dietary scores and patterns across six waves of the Longitudinal Study of Australian Children. British Journal of Nutrition [Internet] 2017 [cited 2026 May 6];117:1137–50. Available from: https://www.cambridge.org/core/journals/british-journal-of-nutrition/article/stability-and-change-in-dietary-scores-and-patterns-across-six-waves-of-the-longitudinal-study-of-australian-children/FBE663C6F7FC6775A57CD170C061C6BE

22. Boggs DA, Palmer JR, Spiegelman D, Stampfer MJ, Adams-Campbell LL, Rosenberg L. Dietary patterns and 14-y weight gain in African American women. Am J Clin Nutr 2011;94:86–94.

23. Agurs-Collins T, Rosenberg L, Makambi K, Palmer JR, Adams-Campbell L. Dietary patterns and breast cancer risk in women participating in the Black Women’s Health Study. Am J Clin Nutr 2009;90:621–8.

24. Nomura SJO, Dash C, Rosenberg L, Yu J, Palmer JR, Adams-Campbell LL. Adherence to diet, physical activity and body weight recommendations and breast cancer incidence in the Black Women’s Health Study. Int J Cancer 2016;139:2738–52.

25. Kumanyika SK, Mauger D, Mitchell DC, Phillips B, Smiciklas-Wright H, Palmer JR. Relative validity of food frequency questionnaire nutrient estimates in the Black Women’s Health Study. Ann Epidemiol 2003;13:111–8.

26. Vávra J, Komárek A, Grün B, Malsiner-Walli G. Clusterwise multivariate regression of mixed-type panel data. Stat Comput [Internet] 2024 [cited 2026 May 6];34:46. Available from: https://link.springer.com/10.1007/s11222-023-10304-5

27. Tan Z, Shen C, Subbarao P, Lou W, Lu Z. A Joint Modeling Approach for Clustering Mixed-Type Multivariate Longitudinal Data: Application to the CHILD Cohort Study. 2022 [cited 2026 May 6]. Available from: https://www.semanticscholar.org/paper/A-Joint-Modeling-Approach-for-Clustering-Mixed-Type-Tan-Shen/734a46b848aa41647952b06a3e60ed59f9f835e3

28. Komárek A, Komárková L. Clustering for multivariate continuous and discrete longitudinal data. The Annals of Applied Statistics [Internet] Institute of Mathematical Statistics; 2013 [cited 2026 May 6];7:177–200. Available from: https://projecteuclid.org/journals/annals-of-applied-statistics/volume-7/issue-1/Clustering-for-multivariate-continuous-and-discrete-longitudinal-data/10.1214/12-AOAS580.full

29. Lu Z, Lou W. Bayesian consensus clustering for multivariate longitudinal data. Stat Med 2022;41:108–27.

30. Lu Z, Chandra NK. A sparse factor model for clustering high-dimensional longitudinal data. Statistics in Medicine [Internet] 2024 [cited 2026 May 6];43:3633–48. Available from: https://onlinelibrary.wiley.com/doi/abs/10.1002/sim.10151

31. Woo JG, Reynolds K, Summer S, Khoury PR, Daniels SR, Kalkwarf HJ. Longitudinal Diet Quality Trajectories Suggest Targets for Diet Improvement in Early Childhood. Journal of the Academy of Nutrition and Dietetics [Internet] 2021 [cited 2026 June 18];121:1273–83. Available from: https://linkinghub.elsevier.com/retrieve/pii/S2212267220312260

32. Talegawkar SA, Jin Y, Xue Q-L, Tanaka T, Simonsick EM, Tucker KL, Ferrucci L. Dietary Pattern Trajectories in Middle Age and Physical Function in Older Age. Newman AB, editor. The Journals of Gerontology: Series A [Internet] 2021 [cited 2026 May 5];76:513–9. Available from: https://academic.oup.com/biomedgerontology/article/76/3/513/5995593

33. Batis C, Mendez MA, Sotres-Alvarez D, Gordon-Larsen P, Popkin B. Dietary pattern trajectories during 15 years of follow-up and HbA1c, insulin resistance and diabetes prevalence among Chinese adults. J Epidemiol Community Health [Internet] 2014 [cited 2026 May 5];68:773–9. Available from: https://jech.bmj.com/lookup/doi/10.1136/jech-2013-203560

34. Wu F, Pahkala K, Juonala M, Rovio SP, Sabin MA, Rönnemaa T, Buscot M-J, Smith KJ, Männistö S, Jula A, et al. Dietary Pattern Trajectories from Youth to Adulthood and Adult Risk of Impaired Fasting Glucose: A 31-year Cohort Study. The Journal of Clinical Endocrinology & Metabolism [Internet] 2021 [cited 2026 June 18];106:e2078–86. Available from: https://academic.oup.com/jcem/article/106/5/e2078/6122544

35. Kerr JA, Gillespie AN, Gasser CE, Mensah FK, Burgner D, Wake M. Childhood dietary trajectories and adolescent cardiovascular phenotypes: Australian community-based longitudinal study. Public Health Nutr [Internet] 2018 [cited 2026 June 18];21:2642–53. Available from: https://www.cambridge.org/core/product/identifier/S1368980018001398/type/journal_article

36. Wright M, Sotres-Alvarez D, Mendez MA, Adair L. The association of trajectories of protein intake and age-specific protein intakes from 2 to 22 years with BMI in early adulthood. Br J Nutr [Internet] 2017 [cited 2026 June 18];117:750–8. Available from: https://www.cambridge.org/core/product/identifier/S0007114517000502/type/journal_article

37. Lee YQ, Colega M, Sugianto R, Lai JS, Godfrey KM, Tan KH, Shek LP-C, Loy SL, Müller Riemenschneider F, Padmapriya N, et al. Tracking of dietary patterns between pregnancy and 6 years post-pregnancy in a multiethnic Asian cohort: the Growing Up in Singapore Towards healthy Outcomes (GUSTO) study. Eur J Nutr [Internet] 2022 [cited 2026 May 5];61:985–1001. Available from: https://link.springer.com/10.1007/s00394-021-02703-z

38. Movassagh E, Baxter-Jones A, Kontulainen S, Whiting S, Vatanparast H. Tracking Dietary Patterns over 20 Years from Childhood through Adolescence into Young Adulthood: The Saskatchewan Pediatric Bone Mineral Accrual Study. Nutrients [Internet] 2017 [cited 2026 May 5];9:990. Available from: https://www.mdpi.com/2072-6643/9/9/990

39. Smithers LG, Golley RK, Mittinty MN, Brazionis L, Northstone K, Emmett P, Lynch JW. Do Dietary Trajectories between Infancy and Toddlerhood Influence IQ in Childhood and Adolescence? Results from a Prospective Birth Cohort Study. Kappen C, editor. PLoS ONE [Internet] 2013 [cited 2026 May 5];8:e58904. Available from: https://dx.plos.org/10.1371/journal.pone.0058904

40. Newby P, Weismayer C, Åkesson A, Tucker KL, Wolk A. Long-Term Stability of Food Patterns Identified by Use of Factor Analysis among Swedish Women. The Journal of Nutrition [Internet] 2006 [cited 2026 May 6];136:626–33. Available from: https://linkinghub.elsevier.com/retrieve/pii/S0022316622081111

