## Supplementary Materials for "Examining stability of dietary patterns across time using repeated measures of dietary intake: Results from Black Women’s Health Study"

**APPENDIX A. DESCRIPTIVE TABLE OF OVERALL CONSUMPTION DISTRIBUTIONS**

**Supplementary Table 1. Frequency distribution of foods observed in BWHS diet assessments (1995, 2001, 2013,2021)**

| **FOOD ITEM** | **cycle** | **< 1 per MONTH** | **1 per MONTH** | **2-3 per MONTH** | **1 per WEEK** | **2 per WEEK** | **3-4 per WEEK** | **≧ 5-6 per WEEK** |
| --- | --- | --- | --- | --- | --- | --- | --- | --- |
| apples pears canned fruit | 1995 | 15.1 | 12.4 | 23.1 | 10.6 | 11.7 | 15.1 | 12 |
|  | 2001 | 10.2 | 14.8 | 24.7 | 6.9 | 25 | 10.6 | 7.8 |
|  | 2013 | 10.4 | 12.9 | 25 | 5.8 | 25.1 | 9.1 | 11.7 |
|  | 2021 | 14.4 | 15.7 | 26.5 | 6.1 | 24.4 | 6.5 | 6.4 |
| bananas | 1995 | 18.4 | 10 | 22.2 | 11.1 | 12.4 | 14.3 | 11.6 |
|  | 2001 | 16.5 | 9.7 | 19.2 | 10 | 14.4 | 17 | 13.3 |
|  | 2013 | 15.8 | 7.9 | 16.3 | 9.7 | 12.7 | 17.4 | 20.2 |
|  | 2021 | 22 | 10 | 19.2 | 8.7 | 12.8 | 15.5 | 11.9 |
| cantaloupe other fruit | 1995 | 35.5 | 16.5 | 18.9 | 10.4 | 7.9 | 6.1 | 4.8 |
|  | 2001 | 6.9 | 22.3 | 22.4 | 13.8 | 20.1 | 9.4 | 5.1 |
|  | 2013 | 5.5 | 17.7 | 18.6 | 13.1 | 24.4 | 12.2 | 8.5 |
|  | 2021 | 8.1 | 23.2 | 18.8 | 12.9 | 22.9 | 9.4 | 4.7 |
| oranges grapefruits | 1995 | 18 | 18 | 25.7 | 5.8 | 17.3 | 8.8 | 6.4 |
|  | 2001 | 18.7 | 18.1 | 22.3 | 10.6 | 10.9 | 10.7 | 8.7 |
|  | 2013 | 21.9 | 15.9 | 21 | 10 | 10.9 | 10.3 | 10 |
|  | 2021 | 26 | 13.8 | 21.2 | 9.1 | 11.4 | 10.7 | 7.8 |
| orange grapefruit juice | 1995 | 10.3 | 6.9 | 13.7 | 9.1 | 10.2 | 15.9 | 33.9 |
|  | 2001 | 13.7 | 13.3 | 12.3 | 8.7 | 10 | 14.8 | 27.2 |
|  | 2013 | 37.5 | 12.1 | 13.9 | 7.5 | 8.2 | 8.5 | 12.2 |
|  | 2021 | 52.9 | 11.4 | 12.1 | 5.9 | 6.2 | 6.7 | 4.9 |
| other fruit drinks | 1995 | 11.7 | 5.4 | 8.5 | 6.3 | 7.9 | 14.2 | 46 |
|  | 2001 | 23.6 | 8.6 | 9.8 | 7.4 | 8.3 | 11.8 | 30.5 |
|  | 2013 | 48.5 | 8.1 | 10.1 | 5.6 | 6.5 | 7.7 | 13.5 |
|  | 2021 | 61.8 | 7.6 | 8.5 | 4.7 | 4.8 | 5.8 | 6.8 |
| beans | 1995 | 18.5 | 18.9 | 27.2 | 14.7 | 20.7 |  |  |
|  | 2001 | 18.1 | 19.1 | 29.8 | 14.9 | 18.2 |  |  |
|  | 2013 | 15.2 | 19.4 | 28.8 | 14 | 22.5 |  |  |
|  | 2021 | 16.5 | 20 | 33.1 | 11.8 | 18.6 |  |  |
| chili with beans | 1995 | 52.9 | 24 | 23.1 |  |  |  |  |
|  | 2001 | 54 | 24.2 | 21.9 |  |  |  |  |
|  | 2013 | 53.1 | 27.4 | 19.5 |  |  |  |  |
|  | 2021 | 53.8 | 27.2 | 19 |  |  |  |  |
| tomatoes | 1995 | 32.7 | 16 | 22.5 | 9.4 | 8.4 | 11 |  |
|  | 2001 | 24.9 | 14.5 | 22.4 | 11.5 | 10.6 | 16 |  |
|  | 2013 | 25.3 | 9.4 | 16.6 | 11 | 14.1 | 23.6 |  |
|  | 2021 | 28.8 | 11.7 | 17 | 9.3 | 12.4 | 20.9 |  |
| broccoli | 1995 | 8.9 | 10.6 | 23.3 | 18 | 14.7 | 24.4 |  |
|  | 2001 | 8.1 | 11.4 | 25.3 | 18.9 | 15.7 | 20.6 |  |
|  | 2013 | 6.5 | 9.9 | 25.7 | 19.1 | 18.5 | 20.3 |  |
|  | 2021 | 8.5 | 11.2 | 27.1 | 16.2 | 17.1 | 19.9 |  |
| spinach | 1995 | 34.9 | 17.2 | 18.7 | 12.1 | 17.2 |  |  |
|  | 2001 | 27.9 | 18.2 | 20.6 | 13.3 | 19.9 |  |  |
|  | 2013 | 12.6 | 11.9 | 20.7 | 15.4 | 39.3 |  |  |
|  | 2021 | 14.4 | 13.9 | 22.1 | 14 | 35.6 |  |  |
| collard greens | 1995 | 26.1 | 24.5 | 27.6 | 10.2 | 11.6 |  |  |
|  | 2001 | 20.4 | 18.1 | 27.3 | 16.6 | 17.7 |  |  |
|  | 2013 | 17.8 | 22.2 | 26.9 | 13.2 | 20 |  |  |
|  | 2021 | 22.4 | 20.8 | 27.5 | 13.3 | 16.1 |  |  |
| cole slaw | 1995 | 34 | 23.1 | 23.2 | 19.7 |  |  |  |
|  | 2001 | 32.6 | 24.8 | 24 | 18.6 |  |  |  |
|  | 2013 | 29.5 | 26.2 | 22 | 22.3 |  |  |  |
|  | 2021 | 30.2 | 25 | 26.1 | 18.7 |  |  |  |
| other vegetables | 1995 | 15.2 | 15.6 | 23 | 14.1 | 11.5 | 20.6 |  |
|  | 2001 | 0.8 | 11.3 | 28.2 | 20 | 25.3 | 14.3 |  |
|  | 2013 | 1.2 | 9.3 | 26.4 | 19.3 | 28.5 | 15.3 |  |
|  | 2021 | 1.5 | 17.2 | 27.7 | 19.8 | 24.1 | 9.7 |  |
| green salad | 1995 | 5.3 | 8.2 | 20.5 | 15.9 | 15.7 | 17.6 | 16.9 |
|  | 2001 | 3.2 | 6.3 | 14 | 13 | 16.9 | 19 | 27.5 |
|  | 2013 | 2.3 | 3.6 | 12.6 | 10.9 | 16.6 | 23.4 | 30.6 |
|  | 2021 | 4.2 | 5.4 | 14.4 | 12.1 | 17.4 | 20.7 | 25.9 |
| fries potatoes | 1995 | 14.3 | 15.3 | 29.1 | 17.3 | 24 |  |  |
|  | 2001 | 14.5 | 18 | 29 | 16 | 22.5 |  |  |
|  | 2013 | 22.2 | 22.8 | 28.2 | 13.8 | 13 |  |  |
|  | 2021 | 21.4 | 22.1 | 31.7 | 12.3 | 12.6 |  |  |
| sweet potatoes | 1995 | 37.3 | 26.1 | 23.1 | 13.5 |  |  |  |
|  | 2001 | 31 | 28.3 | 24.2 | 16.5 |  |  |  |
|  | 2013 | 20.4 | 23.3 | 28.2 | 28.1 |  |  |  |
|  | 2021 | 22.1 | 25.8 | 30.9 | 21.1 |  |  |  |
| other potatoes | 1995 | 10.2 | 17.7 | 31.4 | 40.6 |  |  |  |
|  | 2001 | 13 | 21.3 | 30.8 | 34.9 |  |  |  |
|  | 2013 | 22.5 | 24.4 | 27.9 | 25.3 |  |  |  |
|  | 2021 | 24.4 | 26.8 | 29.6 | 19.1 |  |  |  |
| chips popcorn | 1995 | 13.4 | 12.4 | 22.4 | 15.9 | 12.4 | 11.6 | 12 |
|  | 2001 | 13.1 | 13.1 | 22.4 | 14.9 | 14 | 9.6 | 12.9 |
|  | 2013 | 14.4 | 14.3 | 21.4 | 15.1 | 13.7 | 10.7 | 10.5 |
|  | 2021 | 14.1 | 12.6 | 23.7 | 13.3 | 13.5 | 11.2 | 11.7 |
| high fiber | 1995 | 48.7 | 9.7 | 11.7 | 6.6 | 23.2 |  |  |
|  | 2001 | 42.1 | 11.2 | 12 | 7.1 | 27.6 |  |  |
|  | 2013 | 37.5 | 10.2 | 13 | 7.3 | 32 |  |  |
|  | 2021 | 52.9 | 10.6 | 12.6 | 6.9 | 17.1 |  |  |
| fortified cereals | 1995 | 73.9 | 26.1 |  |  |  |  |  |
|  | 2001 | 70.1 | 29.9 |  |  |  |  |  |
|  | 2013 | 73.7 | 26.3 |  |  |  |  |  |
|  | 2021 | 79 | 21 |  |  |  |  |  |
| other cereals | 1995 | 36.8 | 12.8 | 17.2 | 33.2 |  |  |  |
|  | 2001 | 46.5 | 14.8 | 15 | 23.8 |  |  |  |
|  | 2013 | 67 | 10.3 | 9.8 | 13 |  |  |  |
|  | 2021 | 72.8 | 9.9 | 8.7 | 8.6 |  |  |  |
| other cook cereal | 1995 | 24.7 | 14.5 | 22.7 | 13.1 | 11.2 | 13.8 |  |
|  | 2001 | 20.8 | 15.6 | 23.8 | 13.1 | 12.3 | 14.4 |  |
|  | 2013 | 20.3 | 14.5 | 24 | 12 | 11.6 | 17.6 |  |
|  | 2021 | 20.4 | 13.9 | 24.8 | 11 | 12.5 | 17.4 |  |
| rice | 1995 | 4.6 | 8.3 | 20.8 | 14.9 | 17.4 | 33.9 |  |
|  | 2001 | 5 | 9.1 | 21.8 | 15.9 | 18.6 | 29.5 |  |
|  | 2013 | 9.4 | 14 | 26.3 | 16.7 | 15.7 | 17.9 |  |
|  | 2021 | 12.4 | 15.5 | 30.8 | 13.9 | 14.8 | 12.6 |  |
| spaghetti lasagna pasta | 1995 | 5.5 | 12.7 | 27.6 | 54.3 |  |  |  |
|  | 2001 | 6.6 | 14.9 | 40.6 | 38 |  |  |  |
|  | 2013 | 13.3 | 17.9 | 44.4 | 24.4 |  |  |  |
|  | 2021 | 42.5 | 40.1 | 14.8 | 2.6 |  |  |  |
| white bread | 1995 | 17.7 | 10 | 19.5 | 52.8 |  |  |  |
|  | 2001 | 24.2 | 14.4 | 21.2 | 40.2 |  |  |  |
|  | 2013 | 45.9 | 17.7 | 16.5 | 19.9 |  |  |  |
|  | 2021 | 42.6 | 16.7 | 20.7 | 19.9 |  |  |  |
| dark bread | 1995 | 16.3 | 8.1 | 14.3 | 61.2 |  |  |  |
|  | 2001 | 18.1 | 10.7 | 17.5 | 53.7 |  |  |  |
|  | 2013 | 16.8 | 10.1 | 19.6 | 53.4 |  |  |  |
|  | 2021 | 30 | 12.3 | 22.3 | 35.3 |  |  |  |
| corn bread | 1995 | 24.9 | 22.4 | 27.8 | 24.9 |  |  |  |
|  | 2001 | 27.5 | 24.7 | 26.2 | 21.6 |  |  |  |
|  | 2013 | 33.7 | 27.6 | 22.9 | 15.8 |  |  |  |
|  | 2021 | 38.1 | 28.5 | 22.8 | 10.5 |  |  |  |
| biscuits | 1995 | 20.3 | 19.2 | 27.8 | 32.8 |  |  |  |
|  | 2001 | 26 | 22.4 | 25.3 | 26.3 |  |  |  |
|  | 2013 | 37.9 | 25.2 | 21 | 16 |  |  |  |
|  | 2021 | 43.7 | 22.8 | 21.3 | 12.3 |  |  |  |
| eggs | 1995 | 18.5 | 13.4 | 31.7 | 14.4 | 21.9 |  |  |
|  | 2001 | 14.4 | 11.7 | 29.5 | 16.8 | 27.7 |  |  |
|  | 2013 | 11.8 | 8.4 | 23.9 | 15.6 | 40.3 |  |  |
|  | 2021 | 11.6 | 6 | 21.4 | 14.3 | 46.7 |  |  |
| hamburger | 1995 | 23.2 | 14.7 | 27 | 14.9 | 20.2 |  |  |
|  | 2001 | 25.7 | 15.9 | 24.1 | 15.3 | 19.1 |  |  |
|  | 2013 | 29.8 | 20.6 | 26.5 | 12.3 | 10.8 |  |  |
|  | 2021 | 30.4 | 20.3 | 28.6 | 10.6 | 10.1 |  |  |
| beef | 1995 | 33.1 | 17.9 | 23.6 | 25.3 |  |  |  |
|  | 2001 | 32.3 | 20.4 | 21.9 | 25.4 |  |  |  |
|  | 2013 | 38.9 | 23 | 20.7 | 17.3 |  |  |  |
|  | 2021 | 41 | 23.8 | 22.5 | 12.8 |  |  |  |
| beef stew | 1995 | 63.4 | 19.6 | 16.9 |  |  |  |  |
|  | 2001 | 64.5 | 20.2 | 15.2 |  |  |  |  |
|  | 2013 | 67.9 | 19.7 | 12.4 |  |  |  |  |
|  | 2021 | 69.5 | 19.4 | 11.1 |  |  |  |  |
| liver | 1995 | 79.2 | 20.8 |  |  |  |  |  |
|  | 2001 | 78.3 | 21.7 |  |  |  |  |  |
|  | 2013 | 86.1 | 13.9 |  |  |  |  |  |
|  | 2021 | 86.7 | 13.3 |  |  |  |  |  |
| pork | 1995 | 46.7 | 20.1 | 22.1 | 11 |  |  |  |
|  | 2001 | 38.7 | 22.4 | 23.6 | 15.3 |  |  |  |
|  | 2013 | 43.1 | 22.7 | 22.2 | 12 |  |  |  |
|  | 2021 | 44.4 | 23.8 | 23.2 | 8.6 |  |  |  |
| fried chicken | 1995 | 24.1 | 20.8 | 27.6 | 27.4 |  |  |  |
|  | 2001 | 20.7 | 19.8 | 25.9 | 33.6 |  |  |  |
|  | 2013 | 30.4 | 24.4 | 24.4 | 20.8 |  |  |  |
|  | 2021 | 30.2 | 24.1 | 27 | 18.7 |  |  |  |
| fried fish | 1995 | 37.5 | 23.5 | 22.6 | 16.3 |  |  |  |
|  | 2001 | 33.8 | 26 | 23.7 | 16.5 |  |  |  |
|  | 2013 | 40.1 | 27.6 | 20.1 | 12.2 |  |  |  |
|  | 2021 | 41.4 | 28 | 20.2 | 10.4 |  |  |  |
| other fish | 1995 | 34.2 | 20.3 | 21.6 | 11.3 | 12.6 |  |  |
|  | 2001 | 10.4 | 24.3 | 33.3 | 15.5 | 16.5 |  |  |
|  | 2013 | 8.4 | 15.3 | 29.5 | 18 | 28.8 |  |  |
|  | 2021 | 9.5 | 23.6 | 30.8 | 16.5 | 19.6 |  |  |
| tuna fish | 1995 | 29.6 | 26 | 25 | 19.3 |  |  |  |
|  | 2001 | 25.2 | 27.2 | 27.3 | 20.3 |  |  |  |
|  | 2013 | 31.5 | 25.2 | 24.6 | 18.7 |  |  |  |
|  | 2021 | 35.8 | 27.7 | 24.7 | 11.8 |  |  |  |
| poultry | 1995 | 6.4 | 12.5 | 28.4 | 15.5 | 20.9 | 16.2 |  |
|  | 2001 | 6.8 | 10.1 | 22.8 | 13.1 | 21.5 | 25.8 |  |
|  | 2013 | 9.4 | 9.9 | 23.8 | 13.9 | 22.2 | 20.7 |  |
|  | 2021 | 12.5 | 15.7 | 28.6 | 11.7 | 18.9 | 12.6 |  |
| hot dogs | 1995 | 54.6 | 21.5 | 23.8 |  |  |  |  |
|  | 2001 | 44.2 | 25.7 | 30.1 |  |  |  |  |
|  | 2013 | 52.6 | 25.1 | 22.3 |  |  |  |  |
|  | 2021 | 55.1 | 25 | 19.9 |  |  |  |  |
| ham lunch meats | 1995 | 44.9 | 16 | 19.7 | 19.4 |  |  |  |
|  | 2001 | 45.8 | 18.2 | 18.4 | 17.6 |  |  |  |
|  | 2013 | 54.4 | 17.5 | 15.2 | 12.9 |  |  |  |
|  | 2021 | 57.6 | 16.2 | 15.6 | 10.7 |  |  |  |
| vegetable beef soup | 1995 | 37.7 | 21.7 | 20.4 | 20.3 |  |  |  |
|  | 2001 | 42.8 | 24.5 | 19.3 | 13.4 |  |  |  |
|  | 2013 | 43.2 | 22.4 | 18.5 | 15.9 |  |  |  |
|  | 2021 | 50.7 | 21.4 | 17.5 | 10.4 |  |  |  |
| peanuts peanut butter | 1995 | 43.5 | 21.2 | 19.3 | 16 |  |  |  |
|  | 2001 | 33.9 | 22.6 | 19 | 24.4 |  |  |  |
|  | 2013 | 24.3 | 15.4 | 19.8 | 40.5 |  |  |  |
|  | 2021 | 29.4 | 16.1 | 22.2 | 32.2 |  |  |  |
| bacon sausage | 1995 | 30.9 | 18.2 | 23.1 | 12.5 | 15.2 |  |  |
|  | 2001 | 17.5 | 27.8 | 25.3 | 10.5 | 18.9 |  |  |
|  | 2013 | 16.7 | 23 | 25.8 | 10.9 | 23.6 |  |  |
|  | 2021 | 17.6 | 27 | 22.5 | 10.4 | 22.4 |  |  |
| cheese | 1995 | 21.8 | 12.9 | 24.9 | 11.5 | 12.7 | 16.2 |  |
|  | 2001 | 27 | 14.2 | 19.9 | 11 | 11.1 | 16.7 |  |
|  | 2013 | 30.3 | 11.8 | 16.7 | 11.3 | 12.6 | 17.2 |  |
|  | 2021 | 29.9 | 12.1 | 20.3 | 9.9 | 12.8 | 15 |  |
| yogurt | 1995 | 46.2 | 13.2 | 15.9 | 6.2 | 6.8 | 11.7 |  |
|  | 2001 | 40.2 | 13.9 | 15.5 | 7.1 | 8.2 | 15.2 |  |
|  | 2013 | 25.2 | 9.9 | 13.2 | 7.3 | 11.8 | 32.6 |  |
|  | 2021 | 38.3 | 11.9 | 16.3 | 7.8 | 10.9 | 14.8 |  |
| frozen yogurt | 1995 | 54.5 | 17.4 | 28.2 |  |  |  |  |
|  | 2001 | 67.3 | 16.1 | 16.6 |  |  |  |  |
|  | 2013 | 65.3 | 14.7 | 20 |  |  |  |  |
|  | 2021 | 82 | 10.9 | 7.1 |  |  |  |  |
| whole milk | 1995 | 75.3 | 24.7 |  |  |  |  |  |
|  | 2001 | 67.5 | 32.5 |  |  |  |  |  |
|  | 2013 | 86.9 | 13.1 |  |  |  |  |  |
|  | 2021 | 86.5 | 13.5 |  |  |  |  |  |
| 2% milk | 1995 | 63 | 37 |  |  |  |  |  |
|  | 2001 | 52.9 | 47.1 |  |  |  |  |  |
|  | 2013 | 64.6 | 35.4 |  |  |  |  |  |
|  | 2021 | 71 | 29 |  |  |  |  |  |
| skim milk | 1995 | 68.1 | 31.9 |  |  |  |  |  |
|  | 2001 | 57.2 | 42.8 |  |  |  |  |  |
|  | 2013 | 70.2 | 29.8 |  |  |  |  |  |
|  | 2021 | 83.2 | 16.8 |  |  |  |  |  |
| ice cream | 1995 | 28.6 | 17.9 | 23.4 | 30.1 |  |  |  |
|  | 2001 | 25.4 | 27.3 | 28.4 | 18.9 |  |  |  |
|  | 2013 | 30.8 | 28 | 26.3 | 14.8 |  |  |  |
|  | 2021 | 33.6 | 31 | 23.1 | 12.3 |  |  |  |
| donuts pastries | 1995 | 12.6 | 14.9 | 24.1 | 14.5 | 11.3 | 22.7 |  |
|  | 2001 | 13 | 17.1 | 26.2 | 14.6 | 11.2 | 18 |  |
|  | 2013 | 19.4 | 20.3 | 27.1 | 12.4 | 8.7 | 12.2 |  |
|  | 2021 | 19.1 | 20 | 29.1 | 11.2 | 8.6 | 12.2 |  |
| pies | 1995 | 44.8 | 30.1 | 25.1 |  |  |  |  |
|  | 2001 | 47.1 | 31.3 | 21.6 |  |  |  |  |
|  | 2013 | 54.9 | 28.1 | 17 |  |  |  |  |
|  | 2021 | 61.4 | 26.6 | 12 |  |  |  |  |
| chocolate candy | 1995 | 24.4 | 19.2 | 22.3 | 34.1 |  |  |  |
|  | 2001 | 23.5 | 19.8 | 22.1 | 34.6 |  |  |  |
|  | 2013 | 21.2 | 17.8 | 23.9 | 37.1 |  |  |  |
|  | 2021 | 25.2 | 18.5 | 25.3 | 31 |  |  |  |
| soft drink | 1995 | 28.9 | 20.2 | 13.8 | 37.1 |  |  |  |
|  | 2001 | 20.2 | 24.3 | 25.4 | 30.2 |  |  |  |
|  | 2013 | 42.8 | 26 | 17.5 | 13.7 |  |  |  |
|  | 2021 | 50.2 | 26.2 | 13.8 | 9.8 |  |  |  |
| decaf coffee | 1995 | 74.8 | 25.2 |  |  |  |  |  |
|  | 2001 | 72.7 | 27.3 |  |  |  |  |  |
|  | 2013 | 78.2 | 21.8 |  |  |  |  |  |
|  | 2021 | 81.2 | 18.8 |  |  |  |  |  |
| coffee | 1995 | 53.3 | 46.7 |  |  |  |  |  |
|  | 2001 | 48 | 52 |  |  |  |  |  |
|  | 2013 | 42 | 58 |  |  |  |  |  |
|  | 2021 | 39.9 | 60.1 |  |  |  |  |  |
| tea | 1995 | 19.3 | 20.9 | 13.5 | 15.6 | 30.8 |  |  |
|  | 2001 | 28.2 | 22.7 | 13.7 | 13.5 | 21.9 |  |  |
|  | 2013 | 22.3 | 17.1 | 11.9 | 16.1 | 32.6 |  |  |
|  | 2021 | 26.7 | 18.4 | 12.6 | 14.3 | 28 |  |  |
| red chili sauce | 1995 | 46.1 | 18.6 | 17.1 | 18.2 |  |  |  |
|  | 2001 | 42.3 | 20.4 | 19 | 18.2 |  |  |  |
|  | 2013 | 39.8 | 19.9 | 19.4 | 20.9 |  |  |  |
|  | 2021 | 44.1 | 19.4 | 20.3 | 16.1 |  |  |  |
| salad dressing mayo | 1995 | 8.5 | 9.3 | 23.3 | 14.3 | 16.9 | 27.7 |  |
|  | 2001 | 7 | 13.9 | 20 | 13.7 | 22.5 | 22.9 |  |
|  | 2013 | 9.5 | 9.2 | 20.6 | 13.4 | 23.3 | 24 |  |
|  | 2021 | 11.1 | 18.3 | 21.5 | 14.5 | 20.3 | 14.2 |  |
| butter on bread | 1995 | 51.9 | 13.9 | 14.4 | 19.9 |  |  |  |
|  | 2001 | 34.5 | 17.2 | 19.9 | 28.4 |  |  |  |
|  | 2013 | 41.6 | 17.1 | 16.3 | 25 |  |  |  |
|  | 2021 | 38.6 | 16 | 20.2 | 25.1 |  |  |  |
| margarine on bread | 1995 | 33.3 | 12.9 | 18.1 | 35.8 |  |  |  |
|  | 2001 | 41.5 | 15.3 | 17.1 | 26.1 |  |  |  |
|  | 2013 | 60.7 | 11.5 | 11.2 | 16.6 |  |  |  |
|  | 2021 | 71 | 9.5 | 9.1 | 10.4 |  |  |  |
| milk added to coffee tea | 1995 | 50.1 | 7.8 | 6.7 | 7.1 | 7.4 | 20.9 |  |
|  | 2001 | 46.5 | 9.8 | 7 | 7.3 | 8.9 | 20.6 |  |
|  | 2013 | 43.7 | 7 | 5.5 | 8.1 | 8.7 | 27 |  |
|  | 2021 | 44.5 | 6.6 | 7.2 | 8.2 | 10.8 | 22.7 |  |
| sugar added to coffee tea | 1995 | 36.2 | 9.7 | 9.9 | 10.2 | 10.6 | 23.4 |  |
|  | 2001 | 38 | 12.5 | 9.9 | 8.7 | 11.2 | 19.7 |  |
|  | 2013 | 48.5 | 8.2 | 6.6 | 8.2 | 8 | 20.6 |  |
|  | 2021 | 55.9 | 8.2 | 8.1 | 6.6 | 7.9 | 13.3 |  |

**Supplementary Table 2: Mean and SD at each time cycle for overall and AHEI components.**

|  | **1995** | | **2001** | | **2013** | | **2021** | |
| --- | --- | --- | --- | --- | --- | --- | --- | --- |
| **Scores** | **Mean** | **SD** | **Mean** | **SD** | **Mean** | **SD** | **Mean** | **SD** |
| Overall AHEI | 39.8 | 9.6 | 43.6 | 10.4 | 51.7 | 10.6 | 49.4 | 9.5 |
| Sugar-sweetened beverages | 2.2 | 3.2 | 3.3 | 3.7 | 6.3 | 3.8 | 7.4 | 3.4 |
| Omega-3^+^ | 3.1 | 3.0 | 5.1 | 3.5 | 6.2 | 3.6 | 5.3 | 3.6 |
| PUFA^+^ | 6.6 | 2.2 | 7.4 | 2.0 | 7.8 | 1.9 | 8.2 | 1.8 |
| Red/processed meats | 6.5 | 3.1 | 6.6 | 3.0 | 7.2 | 2.7 | 7.4 | 2.6 |
| Alcohol | 4.0 | 2.7 | 3.9 | 2.5 | 4.5 | 2.7 | 4.5 | 2.8 |
| Nuts^+^ | 2.0 | 2.4 | 2.1 | 2.0 | 3.1 | 3.0 | 2.6 | 2.7 |
| Vegetables^+^ | 2.6 | 2.2 | 2.8 | 2.1 | 3.6 | 2.5 | 2.9 | 2.1 |
| Sodium | 4.9 | 3.2 | 4.8 | 3.2 | 5.0 | 3.3 | 4.7 | 3.1 |
| Trans fats | 4.5 | 1.9 | 4.3 | 1.8 | 4.6 | 1.7 | 4.1 | 1.7 |
| Fruit^+^ | 2.3 | 2.2 | 2.1 | 2.0 | 2.4 | 2.2 | 1.8 | 1.7 |
| Whole grains^+^ | 1.1 | 1.3 | 1.0 | 1.1 | 1.0 | 1.1 | 0.6 | 0.7 |

*Each component reflects an increase in diet quality and consequently lower risk of chronic disease. Positive components where an increase in score reflects an increase in consumption are reflected with a ‘^+^’ superscript. All other components are negative components where an increase in score reflects a decrease in consumption.*

**APPENDIX B. Dietary pattern descriptions at each time point**

**1995 Dietary profiles (K^1995^=5)**

Supplementary Figure 1: Full distribution of 1995 dietary profiles by individual food

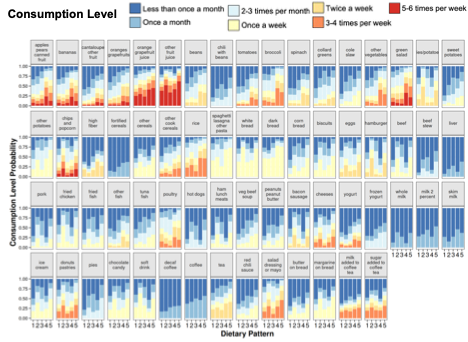

In 1995, participants assigned to profile 1 had the lowest diversity of foods, with only ten foods favoring a positive consumption. Spaghetti, white and dark bread favored consumption once a week Salad dressing, poultry, rice, salad, and broccoli favored consumption at least 2-3 times a month. Participants in profile 2 favored higher consumption frequencies of sweets and snacks such as chips/popcorn (5-6x/week), donuts/pastries (3-4x/week). Hamburger, fries/potatoes, and bacon/sausage favored consumption at twice a week. Participants in profile 3 had a moderate level of consumption (at least 2-3 times per month) over many of the foods queried (43 foods). This included leafy greens (spinach, collard greens, salad), potatoes, rice, pasta, breads, eggs, burger, fried meats, fish, and dessert foods (ice cream, donuts, chocolate) consumed 2-3 times a month. Participants in profile 4 favored more frequent consumption levels of fruits, leafy greens, rice, poultry, other fish. Participants in profile 5 had the largest variety of foods favored to be consumed of all the groups, with higher frequencies of consumption for juice, beans, broccoli, cole slaw, sweet and other potatoes, rice, and sweets (pies, donuts, ice cream).

**2001 Dietary Profiles (K^2001^=6)**

Supplementary Figure 2: Full distribution of 2001 dietary profiles by individual food

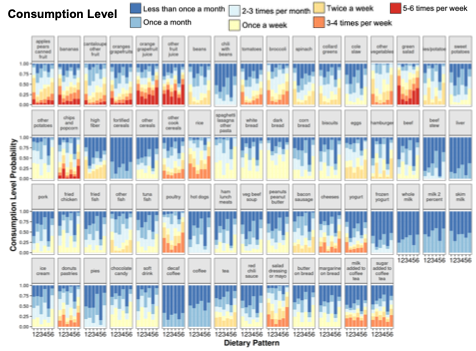

In 2001, participants assigned to profile 1, which had the highest mean AHEI2010 score of 50.8 (SD: 8.7), favored higher frequencies of consumption for fruits (apples, bananas, cantaloupe), vegetables, high fiber, rice, poultry, yogurt, and salad dressing or mayo. Participants assigned to profile 2, which had a mean AHEI2010 score of 39.9 (9.0), favored higher frequencies of consumption of juice, fries, chips and popcorn, cheese, donuts, eggs, and hamburger. Participants in profile 3, with the lowest mean AHEI2010 score of 34.6 (SD: 8.2), had a moderate level of consumption (at least 1-3 times per month) over many of the foods queried. This included fruits (apples bananas – 2-3x/month; cantaloupe, oranges grapefruit – 1/month), vegetables (collard greens, salad, fries/potatoes), carbs (rice, pasta, breads), and meats (poultry, fish, fried chicken, pork). Participants in profiles 4 and 6, with mean AHEI2010 scores of 39.2 (SD:8.2) and 43.8 (SD:7.2) respectively, both favored positive consumption frequency for the same 56 queried foods. However, participants in profile 4 had lower frequencies of consumption compared to those in profile 6. Higher frequencies of consumption for participants in profile 4 were found in foods like juice (at least 5x per week); green salad and salad dressing (twice per week); chocolate, other cereal, bread, fried chicken, and soda (once a week). Participants in profile 6 favored even higher levels of consumption frequencies for fruit, vegetables, chili with beans, fries/potatoes, beef and burgers, bacon/sausage, breads, cheese, added sugar, and donuts. Participants in profile 5, with mean AHEI2010 score 49.1 (SD: 9.7) had the least level of variability amongst frequently consumed foods compared to the other profiles in 2001. Higher levels of consumption frequencies were favored in poultry, rice, broccoli (2-3x per week); green salad (at least 5x per week).

**2013 Dietary Profiles (K^2013^=5)**

Supplementary Figure 3: Full distribution of 2013 dietary profiles by individual food

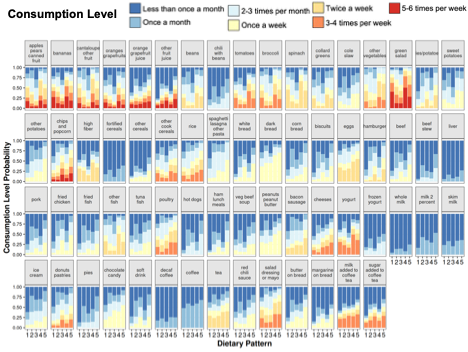

In 2013, participants assigned to profile 1, with a mean AHEI2010 score 58.1 (SD: 8.5), had a low variety of positive consumption favored in their profile pattern, with higher frequencies of consumption for eleven foods. Bananas and green salad favored a consumption of at least 5x a week. Broccoli was favored to be consumed at least 2-3 times a week. Apples/pears, cantaloupe/other fruit, beans, spinach, other vegetables, eggs, and other fish are favored to be consumed twice a week. Dark bread was consumed once a week. Participants assigned to profile 2, with a mean AHEI2010 score 45.5 (SD: 9.3), had a moderate level of consumption (at least 2-3 times per month) over many of the foods queried (42 foods). This included fruits, vegetables, potatoes, rice and pasta at least 2-3 times per month and most meat items for once a month. Participants assigned to profile 3, with a mean AHEI2010 score 49.7 (SD: 7.5), had a moderate level of consumption for most of the 22 foods with positive levels of modal consumption in Figure 1. Higher levels of consumption were noted in bread, eggs, fried chicken, chocolate, and soft drinks. Participants assigned to profile 4, with the highest mean AHEI2010 score 58.6 (SD: 8.0), favored greater levels of consumption for fruits and vegetables, including citrus (orange, grapefruits), tomatoes, and collard greens. Compared to the other profiles, they also were more likely to have greater consumptions of other cooked cereals, poultry, yogurt, salad dressing, beans, and high fiber foods. Participants in profile 5 with the lowest AHEI2010 score 42.6 (SD: 9.1), had the most variety of foods with a level of positive consumption favored in their profile pattern (55 foods). Higher frequency levels of consumption were favored in other fruit juice, cheese, chili with beans, beef stew, vegetable/beef soup, butter on bread, pies, 2% milk, and milk added to coffee/tea, compared to other profiles. Foods favored to be consumed at least once a week included vegetables, potatoes, breads, queries about meat (beef, chicken, fish, liver).

**2021 Dietary Profiles (K^2021^=4)**

Supplementary Figure 4: Full distribution of 2021 dietary profiles by individual food

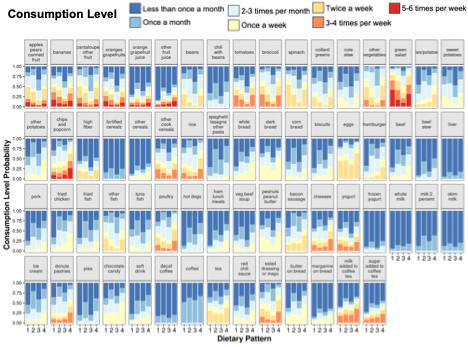

In 2021, coffee and eggs were favored to be consumed at least once a month and at least twice a week, respectively across all four profiles. Participants assigned to profile 1 with the highest AHEI2010 mean score 56.1 (SD: 7.5), had the lowest level of diversity of foods, with a favored positive consumption, compared to the other 2021 profiles. At least twice a week consumption level was found for apples, pears, canned fruit, cantaloupe, beans, spinach, other vegetables, and other fish. Green salad and broccoli that favored higher levels of consumption. Participants assigned to profile 2 with a mean AHEI2010 score 45.6 (SD: 10.1), had a moderate level of consumption for several foods (n=41). These foods were mostly fruit, vegetables, potatoes, beef, chicken, beef, sweets, condiments (mayo, butter). Participants in profile 3 with the lowest AHEI2010 score 45.1 (SD: 8.2), also had low level of diversity of foods with a favored positive consumption for 16 foods. These foods included snack foods (fries/potatoes, chips and popcorn, donuts/pastries) as well as consumption of rice, burgers, fried chicken, bacon/sausage, and broccoli. Participants in profile 4 with a mean AHEI2010 score 49.4 (SD: 8.0), had the highest level of diversity of foods with a favored positive consumption of 45 foods, compared to the other 2021 profiles. Higher consumption was found for fruits, vegetables, breads, meats, and condiments and sauces. Higher levels were favored for salad and typical salad toppings, hamburger, bacon/sausage, peanuts/peanut butter, milk added to coffee, chips and popcorn, bananas, tomatoes, coleslaw.

**APPENDIX C.**

**Supplementary Figure 5: Correlation plots of baseline (1995) derived patterns with future derived patterns. Cells with an ‘X’ over the listed values were not statistically significant (p > 0.05).**

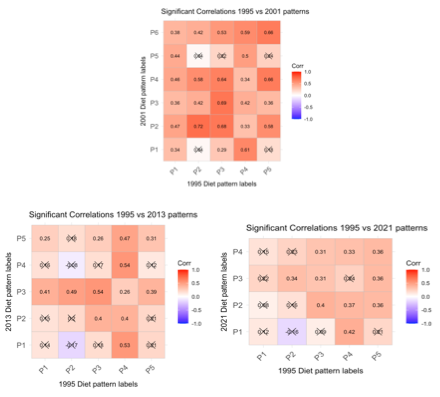

**APPENDIX D. Supplementary Table 3.**Overview of prior diet trajectory approaches.

| Method/Approach | Description | Strengths | Limitations | Examples |
| --- | --- | --- | --- | --- |
| Temporal variation in diet quality adherence (e.g. AHEI, DASH) | Single composite score derived from cumulative intake of aggregated foods/nutrients repeated over time Objective: examine change in score over time | -Standardized  -Comparable across populations/studies -simple to implement  -describes overall trend in diet quality | -different scores penalize foods differently -loss of food-level detail -unreliable for individual variation of food intake and pattern assignment | (10,21,31) |
| Group-based trajectory model | Latent class trajectory individuals in same class share same underlying diet trajectory | -captures distinct group-level trajectories -classification into pattern groups | -within group deviations treated as noise -fixed class membership (individuals can’t switch patterns) | (17,32–34) |
| Growth mixture model | Latent trajectory model with random effects individual variation permitted around group mean pattern | -more flexible than GBTM  -captures heterogeneity around group mean | -fixed class membership  -individuals can’t switch to different patterns | (17,35,36) |
| Pattern stability via principal components or factor analysis (PCA/FA) | Dietary patterns based on shared consumption of similar aggregated food groups and repeated at each time point Stability assessed via correlation and concordance with baseline pattern | -data driven patterns -provide simple numeric measure of pattern similarity over time | -masks which foods drive patterns -limited ability to evaluate long interval stability or individual pattern switching -pattern stability declines with longer time intervals -does not track individual level reassignment to new patterns at new timepoints | (3,37–40) |
